# Impaired type I interferon activity and exacerbated inflammatory responses in severe Covid-19 patients

**DOI:** 10.1101/2020.04.19.20068015

**Authors:** Jérôme Hadjadj, Nader Yatim, Laura Barnabei, Aurélien Corneau, Jeremy Boussier, Hélène Péré, Bruno Charbit, Vincent Bondet, Camille Chenevier-Gobeaux, Paul Breillat, Nicolas Carlier, Rémy Gauzit, Caroline Morbieu, Frédéric Pène, Nathalie Marin, Nicolas Roche, Tali-Anne Szwebel, Nikaïa Smith, Sarah H Merkling, Jean-Marc Treluyer, David Verer, Luc Mouthon, Catherine Blanc, Pierre-Louis Tharaux, Flore Rozenberg, Alain Fischer, Darragh Duffy, Frédéric Rieux-Laucat, Solen Kernéis, Benjamin Terrier

## Abstract

**Background:** Coronavirus disease 2019 (Covid-19) is a major global threat that has already caused more than 100,000 deaths worldwide. It is characterized by distinct patterns of disease progression implying a diverse host immune response. However, the immunological features and molecular mechanisms involved in Covid-19 severity remain so far poorly known.

**Methods:** We performed an integrated immune analysis that included in-depth phenotypical profiling of immune cells, whole-blood transcriptomic and cytokine quantification on a cohort of fifty Covid19 patients with a spectrum of disease severity. All patient were tested 8 to 12 days following first symptoms and in absence of anti-inflammatory therapy.

**Results:** A unique phenotype in severe and critically ill patients was identified. It consists in a profoundly impaired interferon (IFN) type I response characterized by a low interferon production and activity, with consequent downregulation of interferon-stimulated genes. This was associated with a persistent blood virus load and an exacerbated inflammatory response that was partially driven by the transcriptional factor NFĸB. It was also characterized by increased tumor necrosis factor (TNF)-α and interleukin (IL)-6 production and signaling as well as increased innate immune chemokines.

**Conclusion:** We propose that type-I IFN deficiency in the blood is a hallmark of severe Covid-19 and could identify and define a high-risk population. Our study provides a rationale for testing IFN administration combined with adapted anti-inflammatory therapy targeting IL-6 or TNF-α in most severe patients. These data also raise concern for utilization of drugs that interfere with the IFN pathway.

## Introduction

Early clinical descriptions of the first SARS-CoV-2 coronavirus disease (Covid-19) cases at the end of 2019 rapidly highlighted distinct patterns of disease progression^1^. Although most patients experience mild-to-moderate disease, 10 to 20% progress to severe or critical disease, including pneumonia and acute respiratory failure^2^. Based on data from patients with laboratory-confirmed Covid-19 from mainland China, admission to intensive care unit (ICU), invasive mechanical ventilation or death occurred in 6.1%of cases^1^. This proportion of critical cases is higher than that estimated for seasonal Influenza^3^. Additionally, relatively high rates of respiratory failure were reported in young adults (aged 50 years and lower) with previously mild comorbidities (hypertension, diabetes mellitus, overweight)^4^. In severe cases, clinical observations typically describe a two-step disease progression, starting with a mild-to-moderate presentation followed by a secondary respiratory worsening 9 to 12 days after onset of first symptoms^2,5,6^. Respiratory deterioration is concomitant with extension of ground-glass lung opacities on chest computed tomography (CT) scans, lymphocytopenia, high prothrombin time and D-dimer levels^2^. This biphasic evolution marked by a dramatic increase of acute phase reactants in the blood suggests a dysregulated inflammatory host response resulting in an imbalance between pro- and anti-inflammatory mediators. This leads to the subsequent recruitment and accumulation of leukocytes in tissues causing acute respiratory distress syndrome (ARDS)^7^. However, little is known about the immunological features and the molecular mechanisms involved in Covid-19 severity. To test the hypothesis of a virally-driven hyperinflammation leading to severe disease, we employed an integrative approach based on clinical and biological data, in-depth phenotypical analysis of immune cells, standardized whole-blood transcriptomic analysis and cytokine measurements on a group of fifty Covid-19 patients with variable severity from mild to critical.

## Patients and methods

### Study participants

Fifty patients with diagnosis of COVID-19 according to WHO interim guidance and positive SARS-CoV-2 RT-PCR testing on a respiratory sample (nasopharyngeal swab or invasive respiratory sample) were included in this non-interventional study. Inpatients with pre-existing unstable chronic disorders (such as uncontrolled diabetes mellitus, severe obesity defined as body mass index greater than 30, unstable chronic respiratory disease or chronic heart disease) were excluded. Since median duration from onset of symptoms to respiratory failure was previously shown to be 9.5 (interquartile range, 7.0-12.5) days^8^, we analyzed immune signatures between 8 to 12 days after first symptoms for all patients and before the initiation of any anti-inflammatory treatment. Healthy controls were asymptomatic adults, matched with cases on age and with a negative SARS-CoV-2 RT-PCR testing at time of inclusion. The study conforms to the principles outlined in the Declaration of Helsinki, and received approval by the appropriate Institutional Review Board (Cochin-Port Royal Hospital, Paris, France).

The severity of COVID-19 was classified based on the adaptation of the Sixth Revised Trial Version of the Novel Coronavirus Pneumonia Diagnosis and Treatment Guidance. Mild cases were defined as mild clinical symptoms (fever, myalgia, fatigue, diarrhea) and no sign of pneumonia on thoracic computed tomography (CT) scan. Moderate cases were defined as clinical symptoms associated with dyspnea and radiological findings of pneumonia on thoracic CT scan, and requiring a maximum of 3 L/min of oxygen. Severe cases were defined as respiratory distress requiring more than 3 L/min of oxygen and no other organ failure. Critical cases were defined as respiratory failure requiring mechanical ventilation, shock and/or other organ failure necessitating intensive care unit (ICU) cares. Further details of the methods used are provided in the **Supplementary Appendix**.

## Results

### Peripheral blood leukocytes phenotyping

Patients’ characteristics are detailed in the **Supplementary Appendix** and depicted in **Table 1 and Supplementary Figure 1**. As reported in previous studies^9,10^, lymphocytopenia correlates with disease severity (**Figure 1A**). To further characterize it, we used mass cytometry and performed Visualization of t-Distributed Stochastic Neighbor Embedding (viSNE)^11^ to compare cell population densities according to disease severity (**Figure 1B**). viSNE representation and differentiated cell counts showed a decrease in the density of NK cells and CD3+ T cells, including all T cell subsets but more pronounced for CD8+ T cells. This phenotype was more prominent in severe and critical patients, contrasting with an increase in the density of B cells and monocytes (**Figure 1C-F**). No major imbalance in CD4+ and CD8+ T cell naïve/memory subset was observed (**Supplementary Figure 2**). Data on T cell polarization and other minor T cell subsets are indicated in **Supplementary Figure 3**. Plasmablasts were enriched in infected patients (**Figure 1F**), as supported by the increase in genes associated with B cell activation and plasmablast differentiation, such as *IL4R, TNFSF13B* and *XBP1* (**Supplementary Figure 4**).

**Table 1.**
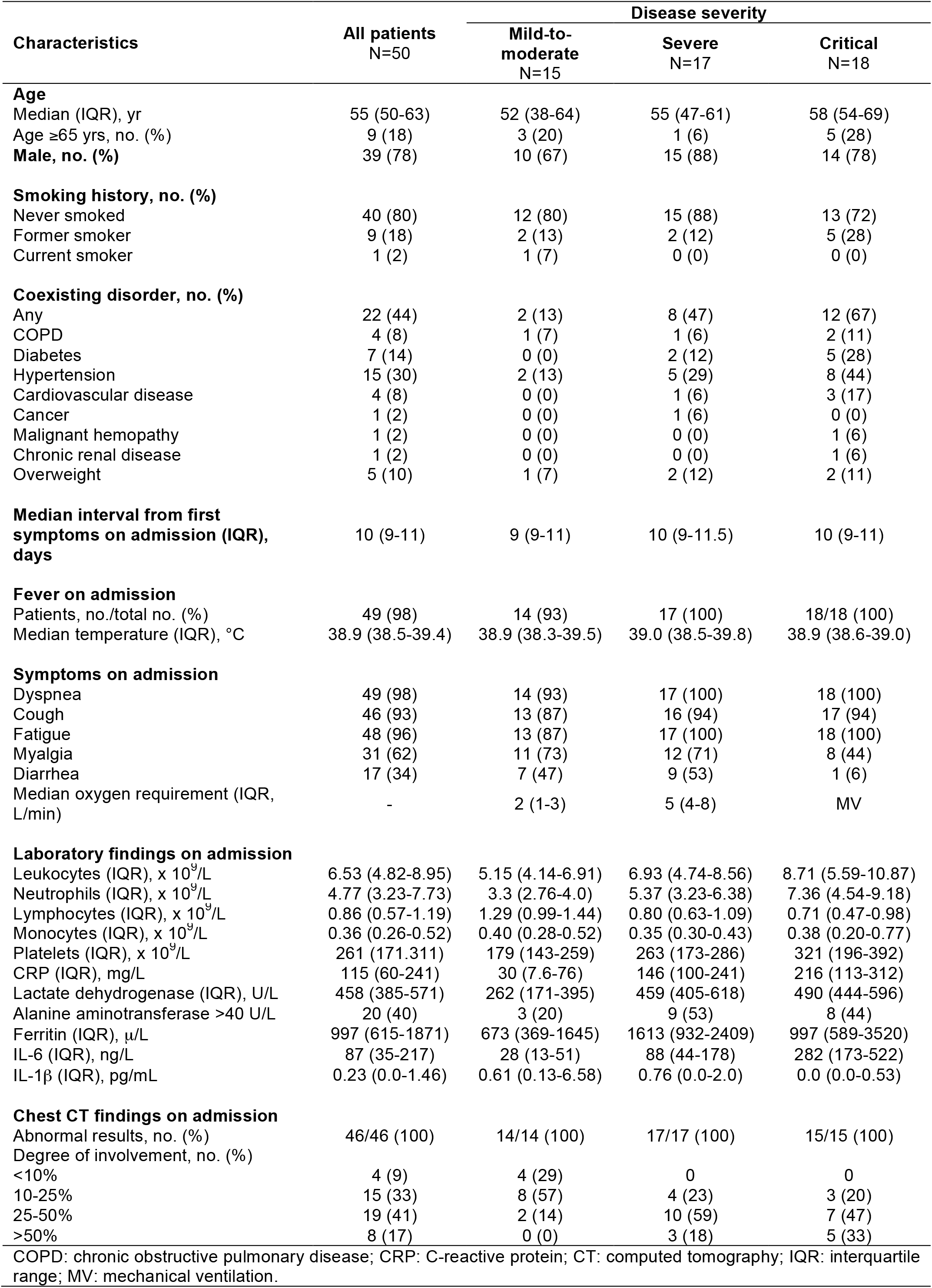
Clinical, laboratory and imaging findings of the study patients.

**Figure 1.**
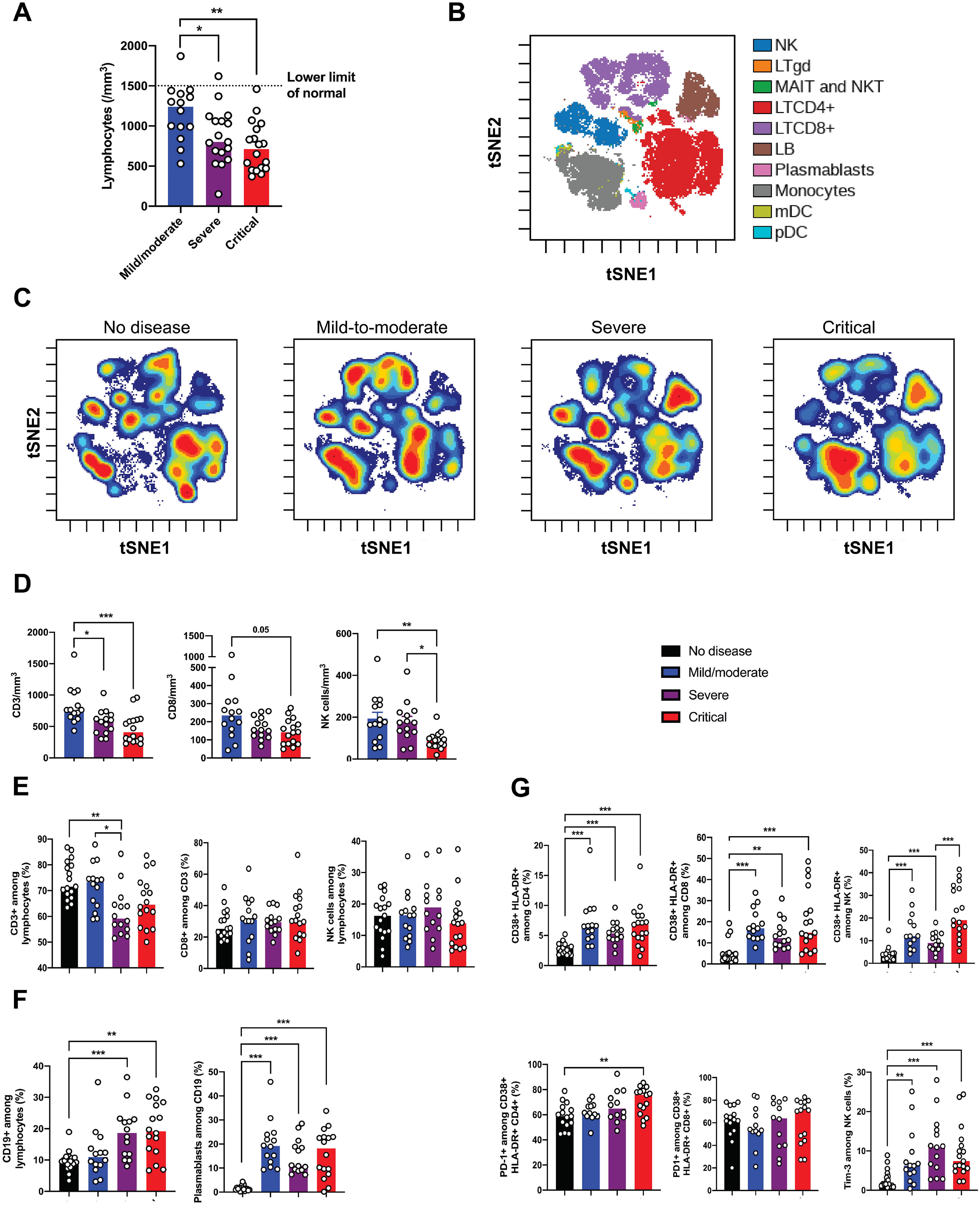
Phenotyping of peripheral blood leukocytes in patients with SARS-CoV-2 infection. **(A)** Lymphocytes count in whole blood from Covid-19 patients was analyzed between days 8 and 12 after onset of first symptoms, according to disease severity. **(B)** viSNE map of blood leukocytes after exclusion of granulocytes, stained with 30 markers and measured with mass cytometry, and automatically separates cells into spatially distinct subsets based on the combination of markers that they express. **(C)** viSNE map colored by cell density across disease severity - no disease, mild-to-moderate, severe and critical. Red color represents the highest density of cells. **(D)** Absolute number of CD3+ T cells, CD8+ T cells and CD3-CD56+ natural killer (NK) cells in peripheral blood from Covid-19 patients, according to disease severity. **(E-F)** Proportions (frequencies) of lymphocyte subsets. Shown are (**E**) proportions of CD3+ T cells among lymphocytes, CD8+ T cells CD3+ among T cells and NK cells among lymphocytes; (**F**) proportions of CD19+ B cells among lymphocytes and CD38hi CD27hi plasmablasts among CD19+ B cells. **(G)** Analysis of the functional status of specific T cell subsets and NK cells based on the expression of activation (CD38, HLA-DR) and exhaustion (PD-1, Tim-3) markers. P values were determined by the Kruskal-Wallis test, followed by Dunn’s post-test for multiple group comparisons with median reported; *P < 0.05; **P < 0.01; ***P < 0.001.

We then assessed the functional status of specific T cell subsets and NK cells using markers of activation (CD25, CD38, HLA-DR) and exhaustion (PD-1, Tim-3) (**Supplementary Figure 5A**). The CD4+ and CD8+ T cell populations were characterized by an increase in CD38+ HLA-DR+ activated T cells in infected patients, with an expression of PD-1 modestly increasing with disease severity (**Figure 1G, Supplementary Figure 5B**). A similar increase in activated NK cells was found in infected patients, especially critical patients, and NK cells displayed a significant increase in Tim-3 expression (**Figure 1G**). Furthermore, expression of exhaustion-related genes, such as *BATF, IRF4* and *CD274*, significantly increased with disease severity (**Supplementary Figure 5C**).

Finally, high annexin-V expression by flow cytometry and upregulation of apoptosis-related genes in the blood from severe and critical patients supported that lymphocytopenia could be partly explained by increased T cell apoptosis (**Supplementary Figure 6**).

### Activation of innate and inflammatory pathways in severe and critical cases

To investigate the immunological transcriptional signatures that characterize disease severity, we quantified the expression of 594 immunology-related genes in blood cells (**Figure 2A**). We identified differentially expressed genes as a function of severity grades (**Figure 2B**). Unsupervised principal component analysis (PCA) separated patients with high disease severity on principal component 1 (PC1), driven by inflammatory and innate immune response encoding genes (GSEA enrichment score with q-value <0.2) (**Figure 2C**). PC2, that was enriched in genes encoding proteins involved in both type I and type II interferon (IFN) responses, distinguished mild-to-moderate patients from the other groups. Collectively, these data suggest a severity grade-dependent increase in activation of innate and inflammatory pathways; in contrast, the IFN response is high in mild-to-moderate patients while it is reduced in more severe patients.

**Figure 2.**
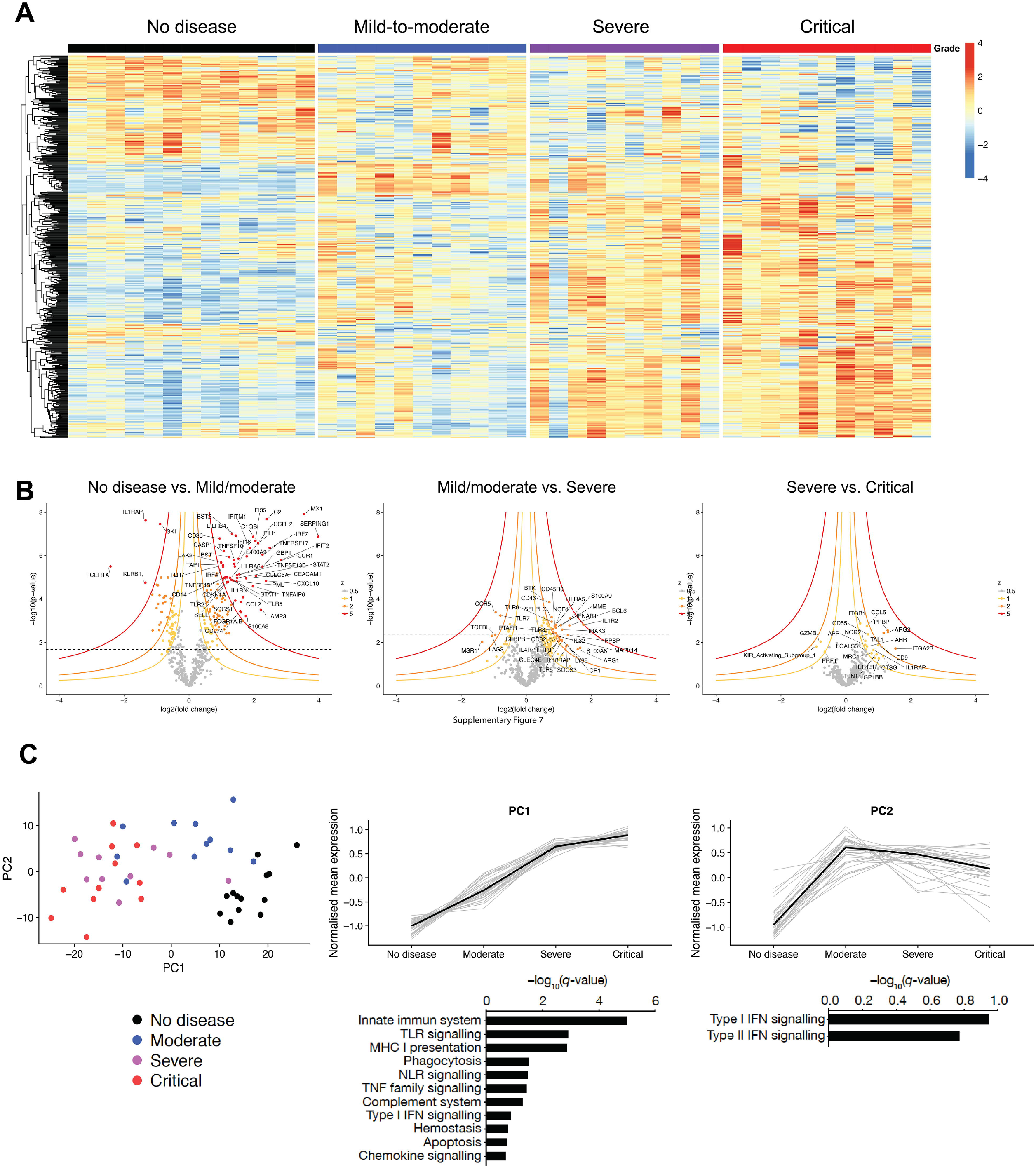
Immunological transcriptional signature of SARS-CoV2 infection. RNA extracted from patient serum and RNA counts of 574 genes were determined by direct probe hybridization using the Nanostring nCounter Human Immunologyv2 kit. (**A**) Heatmap representation of all genes, ordered by hierarchical clustering. No diseases (n=13), mild-to-moderate (n=11), severe (n=10) and critical (n=11). Up-regulated genes are shown in red and down-regulated genes in blue. **(B)** Volcano plots depicting log10(p-value) and log2(fold change), as well as z-value for each group comparison (see Methods). Gene expression comparisons allowed the identification of significantly differentially expressed genes between severity grades (no disease vs mild-moderate, 216 genes; moderate vs severe, 43 genes; severe vs critical, 0 genes). **(C)** *Left panel*, Principal component analysis (PCA) of the transcriptional data. *Middle and right panels*, Kinetics plots showing mean normalized value for each gene and severity grade (each grey line corresponds to one gene). Median values over genes for each severity grade were plotted in black. Gene set enrichment analysis of pathways enriched in PC1 and PC2 are depicted under corresponding kinetic plot.

### Impaired type I interferon antiviral response

Type I IFNs are essential for antiviral immunity^12^. Multiplex gene expression analysis showed an upregulation of genes involved in type I IFN signaling (such as *IFNAR1, JAK1, TYK2*) contrasting with a striking downregulation of interferon-stimulated genes (ISGs) (such as *MX1, IFITM1, IFIT2*) in critical patients (**Figure 3A**). Accordingly, ISG score, based on the expression of 6 ISGs defining a type I IFN signature^13^, was significantly reduced in critical compared to mild-to-moderate patients (**Figure 3B, Supplementary Figure 7A**). Consistently, plasma levels of IFN-α2 protein measured by Simoa digital ELISA^14^ were significantly lower in critical than in mild-to-moderate patients **(Figure 3C)** and correlated with ISG (R^2^=0.27; P=0.0005) (**Supplementary Figure 7B**), while IFN-β was undetectable in all of the patients (data not shown). To further assess the global type I IFN activity, an *in vitro* cytopathic assay was used^15^. IFN activity in serum was significantly lower in severe and critical than in mild-to-moderate patients (**Figure 3D**). Of note, plasmacytoid dendritic cells, the main source of IFN-α^16^, were reduced in infected patients compared to controls, but there was no difference between groups (**Figure 3E**). Finally, we evaluated the response of whole blood cells to an IFN-α stimulation. An increase in ISG score upon IFN-α stimulation was observed, that was similar in infected patients of any severity and controls (**Figure 3F**), indicating that the potential for response to type I IFN was not impacted in critically ill patients. As a possible consequence of impaired IFN-α production, we observed an increased plasma viral load, a possible surrogate marker of uncontrolled lung infection, in severe and critical compared to mild-to-moderate patients, while viral load in nasal swabs was comparable between groups (**Figure 3G**). Overall, these data suggest that patients with severe and critical Covid-19 have an impaired type I IFN production and a lower viral clearance.

**Figure 3.**
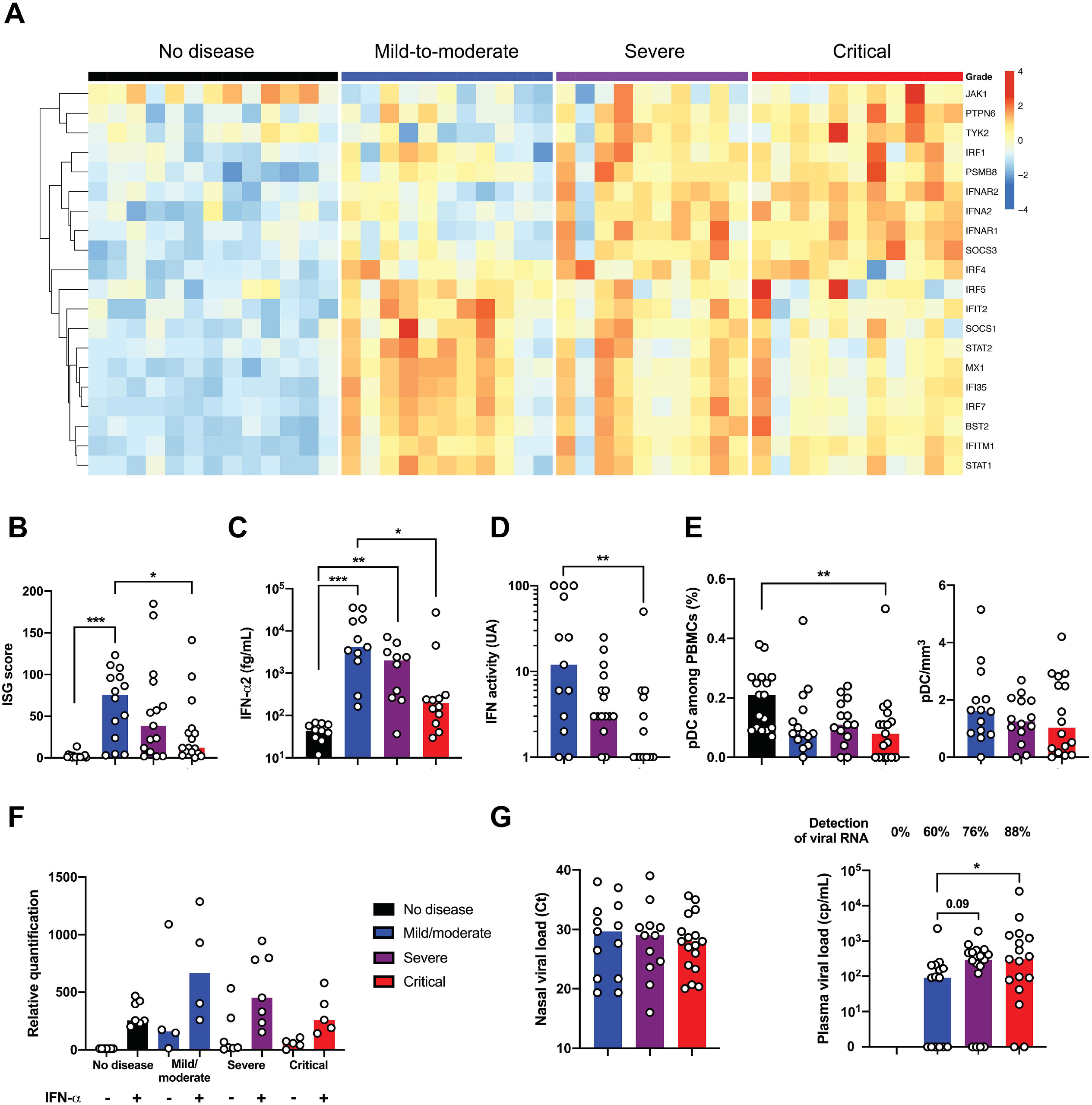
Impaired type I IFN response in severe patients with SARS-CoV2 infection. **(A)** Heatmap showing expression of type 1 IFN-related gene using the reverse transcription- and PCR-free Nanostring nCounter technology in patients with mild-to-moderate (n=11), severe (n=10) and critical (n=11) patients with SARS-CoV2 infection, and 13 healthy controls (no disease). Up-regulated genes are shown in red and down-regulated genes in blue. **(B**) IFN stimulated gene (ISG) score based on expression of 6 genes (*IFI44L, IFI27, RSAD2, SIGLEC1, IFIT1* and *IS15*) measured by q-RT-PCR in whole blood cells from mild-to-moderate (n=14), severe (n=15) and critical (n=17) patients, and 18 healthy controls. **(C)** IFN-α2 (fg/mL) concentration evaluated by SIMOA and **(D)** IFN activity in plasma according to clinical severity. (**E**) Quantification of plasmacytoid dendritic cells (pDC) as a percentage of PBMCs and as cells/ml according to severity group. (**F**) ISG score before and after stimulation of whole blood cells by IFN-α (10^3^UI/mL for 3 hours). **(G)** Viral loads in nasal swab estimated by RT-PCR and expressed in Ct and blood viral load evaluated by digital PCR (**B-F**) Results represent the fold-increased expression compared to the mean of unstimulated controls and are normalized to *GAPDH*. P values were determined by the Kruskal-Wallis test, followed by Dunn’s post-test for multiple group comparisons with median reported; *P < 0.05; **P < 0.01; ***P < 0.001.

### Dissecting the mechanisms of hyperinflammation in severe and critical patients

Severe Covid-19 was reported to induce a cytokine storm^7,17^. Cytokine and chemokine-related genes were found to be increasingly expressed as a function of disease severity in the study cohort (**Figure 4A, Supplementary Figure 8A**). Interestingly, cytokine whole blood RNA levels did not always correlate with protein plasma levels. IL-6, a key player of the cytokine storm in Covid-19^18^, was not detected in peripheral blood at the transcriptional level (**Supplementary Figure 8B**), contrasting with high levels of IL-6 protein (**Figure 4B**). Expression of IL-6-induced genes, such as *IL6R, SOCS3* and *STAT3* were significantly increased (**Supplementary Figure 8B**) reflecting the activation of the IL-6 signaling pathway. TNF-α, a key driver of inflammation, was only moderately upregulated at the transcriptional level (**Supplementary Figure 8C**), whereas circulating TNF-α was significantly increased (**Figure 4C**). Accordingly, TNF pathway-related genes were also upregulated, including *TNFSF10* (**Supplementary Figure 8D-E**), supporting an important role for TNF-α in the induction of inflammation. The discrepancy between RNA quantification and protein measurement suggests that cellular sources of TNF-α and IL-6 originated more likely from the injured lungs and/or endothelial cells. Conversely, while *IL1B* transcripts were significantly upregulated (**Supplementary Figure 8F**), the active form of IL-1β protein was low (**Figure 4D**), suggesting that pro-IL-1β was poorly cleaved and secreted, and that inflammasome activation could not be the main player in the cytokine storm. Circulating IL-1α was also not detected (data not shown). These findings contrast with the detection of high levels of circulating IL-1 receptor antagonist (IL-1RA) and upregulation of *IL1R1* transcripts, indicating an active antagonism of IL-1 in critically ill patients (**Supplementary Figure 8F**). We also detected *IL10* transcripts and IL-10 protein in both severe and critically ill patients (**Figure 4E, Supplementary Figure 8G**). In contrast, no increase of IFN-γ and IL-17A proteins was detected in severe patients (**Supplementary Figure 9**).

**Figure 4.**
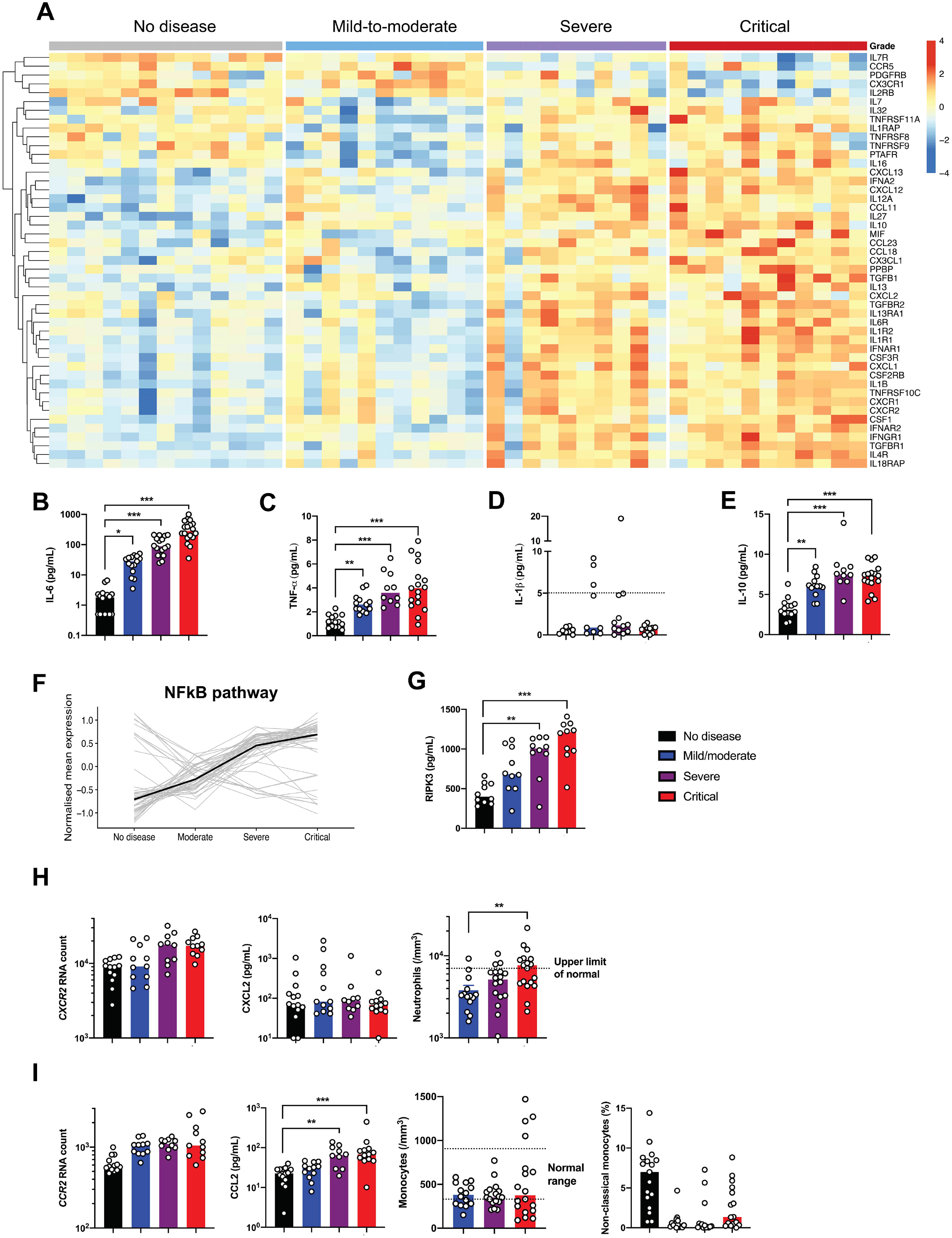
Dissecting the mechanisms of hyperinflammation in severe and critical patients with SARS-CoV2 infection. **(A)** Heatmap showing the expression of cytokines and chemokines that are significantly different in severe and critical patients, and ordered by hierarchical clustering. No diseases (n=13), mild-to-moderate (n=11), severe (n=10) and critical (n=11). Up-regulated genes are shown in red and down-regulated genes in blue. **(B)** Interleukin (IL)-6, **(C)** Tumour necrosis factor (TNF)-α, **(D**) IL-1β and **(E)** IL-10 proteins were quantified in the plasma of patients using simoa technology or a clinical grade ELISA assay (see methods). n= 10 to 18 patients per group. Dashed line depicts the limit of detection (LOD). **(F)** Kinetics plots showing mean normalized value for each gene and severity grade (each grey line corresponds to one gene belonging to the NFĸB pathway). Median values over genes for each severity grade were plotted in black. **(G)** Plasma quantification of receptor-interacting protein kinase (RIPK)-3. n=10 patients per group. **(H)** *left panel*, absolute RNA count for *CXCR2; middle panel*, CXCL2 protein plasma concentration measured by Luminex technology; *right panel*, represents blood neutrophil count depending on severity group. Dashed line depicts the upper normal limit. n= 10 to 13 per group. **(I)** *left panel*, absolute RNA count for *CCR2; second panel*, CCL2 protein plasma concentration measured by Luminex technology; *third panel*, represents blood monocyte count depending on severity group. Dashed lines depict the normal range; *right panel*, represents percentage of non-classical monocytes depending on severity grade. n= 10 to 18 patients per group. RNA data are extracted from the Nanostring nCounter analysis (see methods). P values were determined by the Kruskal-Wallis test, followed by Dunn’s post-test for multiple group comparisons with median reported; *P < 0.05; **P < 0.01; ***P < 0.001.

We explored the expression of transcription factors that may drive this exacerbated inflammation and found that genes specifically upregulated in most of severe and critical ill patients mostly belonged to the NFĸB pathway (**Figure 4F, Supplementary Figure 10**). Aberrant NFĸB activation can result, among several triggering pathways, from excessive innate immune sensors activation by pathogen-associated molecular patterns (PAMPs) (e.g. viral RNA) and/or damage-associated molecular patterns (DAMPs) (e.g. released by necrotic cells and tissue damage). Interestingly, LDH, a marker of necrosis and cellular injury, correlated with disease severity (**Supplementary Figure 1C**), and receptor-interacting protein kinase (RIPK)-3, a key kinase involved in programmed necrosis and inflammatory cell death, was also significantly elevated in severe and critically ill patients (**Figure 4G**) and correlated with LDH (R^2^=0.47; P<0.0001).

The cytokine storm has been associated with massive influx of innate immune cells, namely neutrophils and monocytes, which may aggravate lung injury and precipitate ARDS^19^. We therefore analyzed expression of chemokines and chemokines receptors involved in the trafficking of innate immune cells (**Figure 4A**). While the neutrophil chemokine CXCL2 was detected in the serum but with no difference between groups, its receptor *CXCR2* was significantly upregulated in severe and critical patients (**Figure 4H**). Accordingly, severe disease was accompanied with higher neutrophilia. Monocyte chemotactic factor CCL2 was increased in the blood of infected patients, as well as the transcripts of its receptor *CCR2* and was associated with low circulating inflammatory monocytes (**Figure 4I**), suggesting a role for the CCL2/CCR2 axis in the monocyte chemoattraction into the inflamed lungs. These observations are in accordance with recently published studies in bronchoalveolar fluids from Covid-19 patients, describing the key role of monocytes^19^. Overall, these results support a framework whereby an ongoing inflammatory cascade, initiated by impaired type I IFN production, may be fueled by both PAMPs and DAMPs.

## Discussion

In this study, we identified an impaired type I IFN response associated with high blood viral load in severe and critical Covid-19 patients, that inversely correlated with an excessive NFĸB-driven inflammatory response associated with increased TNF-α and IL-6.

Innate immune sensors, such as TLRs and RIG-I-like receptors, play a key role in controlling RNA virus by sensing viral replication and by alerting the immune system through the expression of a diverse set of antiviral genes^20^. Type I IFNs, which include IFN-α, β and ω, are hence rapidly induced and orchestrate a coordinated antiviral program^21^ via the JAK-STAT signaling pathway and expression of ISGs^21^. We herein observed that most severe Covid-19 patients displayed a lower viral clearance possibly caused by an impaired type I IFN production while cellular response to stimulation was preserved. Further longitudinal studies will be necessary to assess in severe and critical patients whether reduced IFN production is present since the onset of infection, whether the peak is delayed, or whether IFN production is exhausted after an initial peak. SARS-CoV2 may have evolved efficient mechanisms to evade host innate immune pathways. Supporting this hypothesis, it was recently reported that SARS-CoV-2 demonstrated a higher replication rate and a lower induction of host interferon in comparison to SARS-CoV-1^22^. Moreover, virulent human coronaviruses including SARS-CoV-2 have been shown to encode multiple structural and non-structural proteins that antagonize IFN and ISG responses, notably by inhibiting the TANK-binding kinase 1 (TBK1)-dependent phosphorylation and activation of interferon regulatory factor 3 (IRF3), and IFN production^23–26^. Coronaviruses can also inhibit multiple stages of translational initiation^27^. Several hypotheses may be proposed to explain interindividual variability in IFN response to infection. Comorbidities (such as hypertension, diabetes mellitus, overweight) are risk factors for severe Covid-19 and could negatively impact IFN production as well as exacerbate inflammatory responses^28–30^. Genetic host susceptibility can be also suspected since inherited monogenic disorders in children^31,32^ or susceptibility variants in adults^33^, each involving the type I IFN pathway, have been associated with life-threatening influenza infections. Identification of patients with insufficient IFN production could define a high-risk population that could benefit from IFN-α or -β supplementation in conjunction with antiviral drugs when available. Alternatively, IFN-λ (Type III interferon) could be tested as recently proposed^34^, as the receptor is more specifically localized on epithelial cells, which may avoid potential systemic side effects with type I IFN. Conversely, inhibiting IFN production could be deleterious in these patients, reason why agents interfering with the IFN pathway, such as corticosteroids, anti-interferon antibody, JAK1/Tyk2 inhibitors or chloroquine, should be considered with great caution.

Viral replication within the lungs in conjunction with an increased influx of innate immune cells mediates tissue damage and may fuel an auto-amplification inflammatory loop, potentially driven by NFĸB, and ultimately leading to ARDS and respiratory failure. Interestingly, virulent human coronaviruses have been shown to promote multiple form of necrotic cell death such as RIPK3-dependent necroptosis and caspase-1-dependent pyroptosis^35^. These pathways have also been implicated in influenza-induced ARDS in mice and humans^36,37^. Therefore, inflammatory cell death (e.g. necroptosis or pyroptosis) may represent an upstream driver of inflammation that could be targeted^38^.

Our study confirmed that IL-6 plays an important role in pathogenesis and severity of Covid-19. Based on promising case series^39^, randomized trials are currently evaluating the benefits of IL-6 targeted therapy. Our study also provides a case for the inhibition of the TNF axis. Indeed, TNF is highly expressed in alveolar macrophages and anti-TNF does not block immune response in animal models of virus infection^40^. Moreover, TNF blockade induces a significant IL-6 expression reduction^40^. Other targets could also be considered such as chemokines antagonists that block migration of monocytes and neutrophils to the inflamed lungs.

Based on our study, we propose that type I IFN deficiency is a hallmark of severe Covid-19 and infer that severe and critical Covid-19 patients could be potentially relieved from the IFN deficiency by IFN administration and from exacerbated inflammation by adapted anti-inflammatory therapies targeting IL-6 or TNF-α.

## Data Availability

All the microarray data (nanostring) are being processed for upload on GEO database. All the data published are available upon request to the corresponding author.

## Contributors

JH, NY, DD, FRL, SK and BT had the idea for and designed the study and had full access to all of the data in the study and take responsibility for the integrity of the data and the accuracy of the data analysis. JH, NY, AF, DD, FRL, SK and BT drafted the paper. JH, NY, LB, AC, JB, DD, FRL, SK and BT did the analysis, and all authors critically revised the manuscript for important intellectual content and gave final approval for the version to be published. All authors agree to be accountable for all aspects of the work in ensuring that questions related to the accuracy or integrity of any part of the work are appropriately investigated and resolved.

## Acknowledgments

This study was supported by the Fonds IMMUNOV, for Innovation in Immunopathology. The study was also supported by the Institut National de la Santé et de la Recherche Médicale (INSERM), by a government grant managed by the Agence National de la Recherche as part of the “Investment for the Future” program (ANR-10-IAHU-01), and by a grant from the Agence National de la Recherche (ANR-flash Covid19 “AIROCovid” to FRL). J.H. is a recipient of an Institut Imagine MD-PhD fellowship program supported by the Fondation Bettencourt Schueller. L.B. is supported by the EUR G.E.N.E. (reference #ANR-17-EURE-0013) program of the Université de Paris IdEx #ANR-18-IDEX-0001 funded by the French Government through its “Investments for the Future” program

We acknowledge all health-care workers involved in the diagnosis and treatment of patients in Cochin Hospital, especially Célia Azoulay Lauren Beaudeau, Etienne Canoui, Pascal Cohen, Adrien Contejean, Bertrand Dunogué, Didier Journois, Paul Legendre, Jonathan Marey and Alexis Régent. We thank Dr Y. Gaudin for his advices on viral mechanism. We thank all the patients, supporters and our families for their confidence in our work.

